# Mitigating the Severity of COVID-19 Illness in the Primary Care Patient Population through Early Identification and Close Monitoring of Underlying Comorbidities

**DOI:** 10.1101/2022.10.07.22280827

**Authors:** Payal Parikh, Patricia Greenberg, Shmuel Halpert, Allison Abrishami, Liora Rabizadeh, Hannah Shayefar, Lauren Tetelbaun, Dovid Friedman, Jeffrey Kaminetzky

## Abstract

**Purpose:** Prior studies have identified risk factors which prognosticate severity of SARS-CoV-2 illness among hospitalized patients. Since the majority of patients first present to ambulatory care sites, there is a need to identify early predictors of disease progression in this population.

**Methods:** This retrospective cohort study investigated the impact of underlying comorbid conditions on SARS-CoV-2 infection severity in the ambulatory setting. All patients who presented to a single federally qualified health center (FQHC) between March-May 2020 with a positive SARS-CoV-2 test were reviewed for inclusion. Patient demographics, symptomology, prior medical history, and outcomes were collected.

**Results:** 301 patients were included, with nearly equal numbers of patients with (n=151) and without (n=150) underlying comorbidities. Overall, 269 patients (89%) had a mild outcome and 32 patients (11%) had a severe outcome. Advanced age (OR: 9.4 [95% CI: 3.4-27.4], p < 0.001) and male gender (OR: 3.2 [95% CI: 1.2-9.8], p = 0.02) were significant predictors of severe outcomes. Additionally, every obesity category (1: BMI = 30.0–34.9; 2: BMI = 35–39.9; 3: BMI = 40.0+) was associated with more severe outcomes compared to non-obese (OR: 3.5, p = 0.05; OR: 5.2, p = 0.03; OR: 13.9, p = 0.01). Compared to an HbA1C < 6, an HbA1C of 7.1–8.0 showed a clinically significant association.

**Conclusion:** SARS-CoV-2 severity can be prognosticated in the ambulatory population by the presence and severity of pre-existing comorbidities. Early identification and risk stratification of these comorbidities will allow clinicians to develop plans for closer monitoring to prevent severe illness.

## Introduction

Patients infected with SARS-CoV-2 present with variable signs and symptoms, from asymptomatic to Acute Respiratory Distress Syndrome (ARDS)^1^. The relationship between severity of illness and nonmodifiable risk factors is well established. However, modifiable risk factors, such as underlying comorbid conditions are presently under investigation. In hospitalized patients, poor prognosis is linked with underlying comorbidities including cardiovascular disease, immunodeficiency, lung disease, and the components of the metabolic syndrome (obesity, hypertension, diabetes, and hyperlipidemia)^**2**^. In addition, patients with diabetes have increased rates of hospital and ICU admissions with increasing morbidity and mortality^2,3^ and patients with COPD are four times more likely to contract SARS-CoV-2, with a higher likelihood of developing more severe illness compared to patients without COPD^4^. While disease progression has been extensively evaluated in the inpatient population, there is little evidence predicting disease progression in primary care patients who present to the outpatient setting. As such, the ability to prognosticate the potential severity of illness and develop a plan for more frequent monitoring is necessary.

This study reports on the relationship between underlying comorbidities and disease progression on a retrospective cohort of SARS-CoV-2 positive primary care patients. The goal is to guide ambulatory physicians’ focus on more frequent monitoring of these high-risk patients, thus allowing for earlier and more aggressive intervention.

## Methods

### Participants

This retrospective cohort study utilized patient chart data from the ambulatory population presenting to the CHEMED Health Center in Lakewood, New Jersey. Records for all adult patients aged 18 or older, who received SARS-CoV-2 testing at the health center between March 1 - May 31, 2020 were reviewed for study inclusion. All SARS-CoV-2 positive patients (confirmed by PCR testing or significant levels of IgG antibodies) with underlying comorbidities - defined as any patient who had a prior or current diagnosis of diabetes, hypertension, obesity, and/or a history of previous thromboembolic event - were identified and selected for analysis. Confirmed SARS-CoV-2 positive controls (those without underlying comorbidities) were randomly selected and matched to those with comorbidities, based on the combination of three demographic characteristics: age group (1: 18-40 years old, 2: 41-64, 3: 65-80, 4: 81+), sex (female or male), and self-selected ethnicity, which was pared down to the NIH recommended racial and ethnic categories^5^. The randomized matching was performed using Microsoft Excel, first matching on the combination of age group and sex, and then on race as a tertiary factor. Study exclusion criteria included age below 18 and pregnancy at the time of health center visit.

### Study Design

#### Medical Record Review and Comorbidity Classification

Retrospective chart reviews were conducted using the CHEMED EMR database for all participants in the study. Chart extraction included self-selected ethnicity, comorbid conditions, current medication list, and recent BMI calculation, blood pressure measurement, and HbA1c level, when available. The comorbidities were defined on a scale of 1-3 as follows: Diabetes by HbA1c (1: <6.4; 2: 6.5-8.9; 3: >9) based on ADA Guidelines^6^, Hypertension by blood pressure (1: 120-129/<80, 2: 130-139/80-89, 3: >=140/>=90) based on the JNC8 Guidelines^7^, the presence of anti-hypertensive medications, specifically ACE Inhibitors or Angiotensin Receptor Blockers (ARBs), and Obesity by body mass index (BMI) (1: 30-34.9, 2: 35-39.9, 3: >=40). History of prior thromboembolic events was categorized as “yes” or “no” via an absolute count. IRB approval was obtained through the primary author’s institutional requirements.

#### Procedure and Telephone Survey

As part of routine care, all patients who tested positive for SARS-CoV-2 were contacted regarding their health status using a follow-up questionnaire which included the patient’s health status post SARS-CoV-2 infection, initial presenting symptoms, and severity of illness including the need for home oxygen, and/or hospitalization (Supplemental Figure 1). Past medical history was confirmed as part of routine care. For non-English speaking patients, a translator line was used. Researchers ranked the patients’ SARS-CoV-2 infection severity on a scale from 1 to 4 using a modified version of the NIH Clinical Spectrum of SARS-CoV-2 infection tool^8^ (1: asymptomatic; 2: mildly symptomatic; 3: symptomatic and requiring oxygen (moderate), 4: hospitalized (severe/critical)).

To align with the initial ambulatory diagnostic process, disease severity was further pared down to, mild and severe; differentiated by the need for additional therapies. Mild was defined by either asymptomatic or symptomatic presentation alone, while severe was defined by symptomatic presentation requiring supplemental oxygen or hospitalization.

#### Statistical Analysis

In order to examine balance in the charts selected for analysis, all study measures were first summarized overall and by their comorbidity group status using range, mean and standard deviation (SD), and median with interquartile range (IQR) for continuous measures, frequency with percentage for the categorical measures, and a combination of frequency with percentage and median with IQR for ordinal measures. Bivariate group comparisons were done using Pearson Chi-Square, Fisher’s Exact, or Wilcoxon Rank Sum tests, where appropriate.

The primary outcome, SARS-CoV-2 severity, was explored with both bivariate and multivariate methods. In the bivariate analyses, the SARS-CoV-2 severity outcome was compared across the study measure-based groups using Kruskal-Wallis, Wilcoxon Rank Sum, Pearson Chi-Square, and Fisher’s Exact tests for association, and spearman correlation testing for ordinal vs. ordinal or ordinal vs. non-normally distributed continuous measures. Multivariable Firth logistic regression models were then used to examine which combination of study measures were predictive of the 2 level (“Mild”, “Severe”) SARS-CoV-2 severity outcome. Patient demographics and comorbidities were used as the primary predictors for these models. Results of the models are presented are presented as coefficient estimates and as odds ratios (OR) with 95% confidence intervals (CI).

All statistical analyses were completed using R version 4.1.1 statistical software^9^, including the use of the “DescTools” and “logistf” packages^10,11^, and any two-sided p-value < 0.05 was considered to be statistically significant.

## Results

We analyzed clinical data from a total of 301 patients diagnosed with SARS-CoV-2, with a nearly equal subset of patients with underlying comorbidities (n=151) and those without (n=150). While gender was well matched between the groups (p = 0.59), there was a significant difference in the 4-level age categorization (p = 0.01); therefore, age was instead collapsed into a 2-level categorization (1: 18-64 vs 2: 65+) as shown in Table 1, which was then balanced between the groups (p = 0.35). The patient population was predominantly White or Caucasian race (97%), which was also well-matched among the two arms (p = 0.42). However, when combined with Hispanic/Latino ethnicity, there was a much higher proportion of Non-Hispanic White patients in the control group compared to the comorbidity group (96.0% vs 80.8%, p < 0.001).

**Table 1:** Study patient demographics overall and by group.

| **Study Measure** | **Total Study Sample**<br>**(n = 301)** | **Group 1:**<br>**Comorbidities**<br>**(n = 151)** | **Group 2:**<br>**Controls**<br>**(n = 150)** | **P-Value** |
| --- | --- | --- | --- | --- |
| **Age Category (4-level), n (%)** |  |  |  | p = 0.01 |
| 18-40 years old (1) | 119 (39.5) | 45 (29.8) | 74 (49.3) |  |
| 41-64 years old (2) | 133 (44.2) | 78 (51.6) | 55 (36.7) |  |
| 65-80 years old (3) | 40 (13.3) | 22 (14.6) | 18 (12.0) |  |
| 81+ years old (4) | 9 (3.0) | 6 (4.0) | 3 (2.0) |  |
| **Age Category (2-level), n (%)** |  |  |  | p = 0.35 |
| 18-64 years old (1) | 252 (83.7) | 123 (81.5) | 129 (86.0) |  |
| 65+ years old (2) | 49 (16.3) | 28 (18.5) | 21 (14.0) |  |
| **Gender, n (%)** |  |  |  | p = 0.59 |
| Female | 117 (38.9) | 61 (40.4) | 56 (37.3) |  |
| Male | 184 (61.1) | 90 (59.6) | 94 (62.7) |  |
| **Race, n (%)** |  |  |  | p = 0.42 |
| Asian (1) | 4 (1.3) | 2 (1.3) | 2 (1.3) |  |
| Black or African American (2) | 5 (1.7) | 4 (2.6) | 1 (0.7) |  |
| Mexican American Indian (3) | 1 (0.3) | 1 (0.7) | 0 (0.0) |  |
| White or Caucasian (1) | 291 (96.7) | 144 (95.4) | 147 (98.0) |  |
| **Ethnicity, n (%)** |  |  |  | p < 0.001 |
| Hispanic or Latino (1) | 28 (9.3) | 25 (16.6) | 3 (2.0) |  |
| Not Hispanic or Latino (2) | 271 (90.0) | 124 (82.1) | 147 (98.0) |  |
| Refused to Report (9) | 2 (0.7) | 2 (1.3) | 0 (0.0) |  |
| **Race + Ethnicity, n (%)** |  |  |  | p < 0.001 |
| Asian (1) | 2 (0.7) | 0 (0.0) | 2 (1.3) |  |
| Hispanic (2) | 25 (8.3) | 22 (14.6) | 3 (2.0) |  |
| Mixed Race/Ethnicity (3) | 3 (1.0) | 3 (2.0) | 0 (0.0) |  |
| Non-Hispanic Black (4) | 5 (1.7) | 4 (2.6) | 1 (0.7) |  |
| Non-Hispanic White (5) | 266 (88.4) | 122 (80.8) | 144 (96.0) |  |

As shown in Table 2, underlying health conditions in the comorbidity group (n=151) were obesity 76.2%, hypertension 46.4%, diabetes 27.2%, asthma 7.3%, current smoker 6.0%, COPD 0.7%, and no patients were identified with having a history of thromboembolic events. The mean (SD) conditions per patient in the comorbidity arm was 1.6 (0.7) and the median (IQR) was 2 [1, 2].

**Table 2:** Patient comorbidity details.

| **Comorbidity Measure** | **Comorbidity Group**<br>**(n = 151)** |
| --- | --- |
| **Diabetes, n (%)** |  |
| No (0) | 110 (72.8) |
| Yes (1) | 41 (27.2) |
| **Diabetic Category, n (%)** | n = 41 |
| HbA1C < 6.5 (1) | 14 (34.1) |
| HbA1C < 7 (2) | 8 (19.5) |
| HbA1C = 7.1 - 8.0 (3) | 4 (9.8) |
| HbA1C = 8.1 - 9.0 (4) | 6 (14.6) |
| HbA1C > 9.0 (5) | 6 (14.6) |
| Unknown (Blank) | 3 (7.3) |
| **History of Hypertension, n (%)** |  |
| No (0) | 81 (53.6) |
| Yes (1) | 70 (46.4) |
| **Hypertension Category at Visit, n (%)** |  |
| Normal: < 120 (0) | 32 (21.2) |
| Elevated: 120-129 (1) | 11 (7.3) |
| HTN Stage 1: 130-139/80-89 (2) | 44 (29.1) |
| HTN Stage 2: 140+/90+ (3) | 64 (42.4) |
| **ACE-I or ARB at Visit, n (%)** |  |
| No | 100 (66.2) |
| Yes: |  |
| ACE-I | 24 (15.9) |
| ARB | 9 (6.0) |
| ARB & ACE-I | 5 (3.3) |
| Unknown | 13 (8.6) |
| **Obesity, n (%)** |  |
| No (0) | 36 (23.8) |
| Yes (1) | 115 (76.2) |
| **Obesity Category, n (%)** | n = 115 |
| BMI = 30.0 - 34.9 (1) | 66 (57.4) |
| BMI = 35.0 - 39.9 (2) | 34 (29.6) |
| BMI = 40.0+ (3) | 15 (13.0) |
| **Current Smoker, n (%)** |  |
| No (0) | 142 (94.0) |
| Yes (1) | 9 (6.0) |
| **Asthma, n (%)** |  |
| No (0) | 140 (92.7) |
| Yes (1) | 11 (7.3) |
| **COPD, n (%)** |  |
| No (0) | 150 (99.3) |
| Yes (1) | 1 (0.7) |
| **Use of Inhaler for Asthma/COPD, n (%)** |  |
| No (0) | 142 (94) |
| Yes (1) | 7 (4.6) |
| Unknown | 2 (1.3) |
| **History of Blood Clots, n (%)** |  |
| No (0) | 151 (100.0) |
| Yes (1) | 0 (0.0) |
| **Total Number of Comorbidities** |  |
| Mean (SD) | 1.6 (0.7) |
| Median [Q1, Q3] | 2 [1, 2] |

Per Table 3, the majority (87.7%) of patients were symptomatic at presentation during SARS-CoV-2 infection diagnosis. In the total study population, more than 50% of the patients presented with fever, weakness and fatigue, and malaise. More patients presented with cough, shortness of breath, and cardiopulmonary symptoms in the comorbidity group while, more patients presented with weakness and fatigue in the control group. For progression of illness, the majority of patients in both the comorbidity (84.1%) and control (94.7%) groups were identified as symptomatic without the need for additional therapies. A significantly higher proportion of patients in the comorbidity group versus the control group presented with a higher severity of illness, defined as symptomatic requiring home oxygen or subsequent hospitalization (15.9% vs 5.3%, p = 0.003), respectively.

**Table 3:** Patient COVID-19 characteristics and outcomes overall and by group.

| **Study Measure or Outcome** | **Total Study Sample**<br>**(n = 301)** | **Group 1:**<br>**Comorbidities**<br>**(n = 151)** | **Group 2:**<br>**Controls**<br>**(n = 150)** | **P-Value** |
| --- | --- | --- | --- | --- |
| **COVID Related Measures** |  |  |  |  |
| **Symptomatic, n (%)** |  |  |  | p = 0.61 |
| No (0) | 37 (12.3) | 20 (13.2) | 17 (11.3) |  |
| Yes (1) | 264 (87.7) | 131 (86.8) | 133 (88.7) |  |
| **Cough/Shortness of Breath, n (%)** |  |  |  | p = 0.09 |
| No (0) | 160 (53.2) | 73 (48.3) | 87 (58.0) |  |
| Yes (1) | 141 (46.8) | 78 (51.7) | 63 (42.0) |  |
| **Shortness of Breath, n (%)** |  |  |  | p = 0.046 |
| No (0) | 241 (80.1) | 114 (75.5) | 127 (84.7) |  |
| Yes (1) | 60 (19.9) | 37 (24.5) | 23 (15.3) |  |
| **Diarrhea, n (%)** |  |  |  | p = 0.01 |
| No (0) | 288 (95.7) | 140 (92.7) | 148 (98.7) |  |
| Yes (1) | 13 (4.3) | 11 (7.3) | 2 (1.3) |  |
| **Weakness/Fatigue, n (%)** |  |  |  | p = 0.003 |
| No (0) | 142 (47.2) | 84 (55.6) | 58 (38.7) |  |
| Yes (1) | 159 (52.8) | 67 (44.4) | 92 (61.3) |  |
| **Fatigue, n (%)** |  |  |  | p = 0.44 |
| No (0) | 203 (67.4) | 105 (69.5) | 98 (65.3) |  |
| Yes (1) | 98 (32.6) | 46 (30.5) | 52 (34.7) |  |
| **Fever, n (%)** |  |  |  | p = 0.60 |
| No (0) | 137 (45.5) | 71 (47.0) | 66 (44.0) |  |
| Yes (1) | 164 (54.5) | 80 (53.0) | 84 (56.0) |  |
| **Loss of Smell, n (%)** |  |  |  | p = 0.03 |
| No (0) | 193 (64.1) | 106 (70.2) | 87 (58.0) |  |
| Yes (1) | 108 (35.9) | 45 (29.8) | 63 (42.0) |  |
| **Cardiopulmonary Symptom(s), n (%)** |  |  |  | p = 0.09 |
| No (0) | 154 (51.2) | 70 (46.4) | 84 (56.0) |  |
| Yes (1) | 147 (48.8) | 81 (53.6) | 66 (44.0) |  |
| **GI Symptom(s), n (%)** |  |  |  | p < 0.001 |
| No (0) | 269 (89.4) | 122 (80.8) | 147 (98.0) |  |
| Yes (1) | 32 (10.6) | 29 (19.2) | 3 (2.0) |  |
| **Constitutional Symptom(s), n (%)** |  |  |  | p = 0.06 |
| No (0) | 112 (37.2) | 64 (42.4) | 48 (32.0) |  |
| Yes (1) | 189 (62.8) | 87 (57.6) | 102 (68.0) |  |
| **Musculoskeletal Symptom(s), n (%)** |  |  |  | p = 0.63 |
| No (0) | 248 (82.4) | 126 (83.4) | 122 (81.3) |  |
| Yes (1) | 53 (17.6) | 25 (16.6) | 28 (18.7) |  |
| **Vision Symptom(s), n (%)** |  |  |  | p = 0.37 |
| No (0) | 297 (98.7) | 148 (98.0) | 149 (99.3) |  |
| Yes (1) | 4 (1.3) | 3 (2.0) | 1 (0.7) |  |
| **Brain Fog/Confusion Symptom(s), n (%)** |  |  |  | p = 1.0 |
| No (0) | 298 (99.0) | 149 (98.7) | 149 (99.3) |  |
| Yes (1) | 3 (1.0) | 2 (1.3) | 1 (0.7) |  |
| **"Other" Symptom(s), n (%)** |  |  |  | p = 0.02 |
| No (0) | 184 (61.1) | 82 (54.3) | 102 (68.0) |  |
| Yes (1) | 117 (38.9) | 69 (45.7) | 48 (32.0) |  |
| **Total Number of Symptoms** |  |  |  | p = 0.58 |
| Range | 0 - 9 | 0 - 9 | 0 - 6 |  |
| Mean (SD) | 2.6 (1.7) | 2.7 (1.9) | 2.5 (1.5) |  |
| Median [Q1, Q3] | 3 [1, 4] | 3 [1, 4] | 3 [2, 4] |  |
| **Patient Care & Outcome Measures** |  |  |  |  |
| **Sent Home from Doctor on O2, n (%)** |  |  |  | p = 1.0 |
| No (0) | 295 (98.0) | 148 (98.0) | 147 (98.0) |  |
| Yes (1) | 6 (2.0) | 3 (2.0) | 3 (2.0) |  |
| **Admitted to Hospital, n (%)** |  |  |  | p = 0.002 |
| No (0) | 273 (90.7) | 129 (85.4) | 144 (96.0) |  |
| Yes (1) | 28 (9.3) | 22 (14.6) | 6 (4.0) |  |
| **Sent to ICU, n (%)** |  |  |  | p = 0.02 |
| No (0) | 291 (96.7) | 142 (94.0) | 149 (99.3) |  |
| Yes (1) | 10 (3.3) | 9 (6.0) | 1 (0.7) |  |
| **Ventilation Needed, n (%)** |  |  |  | p = 0.21 |
| No (0) | 295 (98.0) | 146 (96.7) | 149 (99.3) |  |
| Yes (1) | 6 (2.0) | 5 (3.3) | 1 (0.7) |  |
| **Sent Home from Hospital on O2, n (%)** |  |  |  | p = 0.72 |
| No (0) | 293 (97.3) | 146 (96.7) | 147 (98.0) |  |
| Yes (1) | 8 (2.7) | 5 (3.3) | 3 (2.0) |  |
| **Severity of Case (4-level), n (%)** |  |  |  | p = 0.18 |
| Asymptomatic infection (1) | 35 (11.6) | 20 (13.2) | 15 (10.0) |  |
| Symptomatic but not requiring anything (2) | 234 (77.8) | 107 (70.9) | 127 (84.7) |  |
| Symptomatic but requiring home oxygen (3) | 4 (1.3) | 2 (1.3) | 2 (1.3) |  |
| Symptomatic requiring hospitalization (4) | 28 (9.3) | 22 (14.6) | 6 (4.0) |  |

When collapsing the severity of SARS-CoV-2 illness outcome to 2 levels: “Mild” (defined as being either asymptomatic or having symptoms, but not requiring additional therapy) versus “Severe” (defined as having symptoms and requiring at home oxygen or hospitalization), there were 269 patients who had a Mild outcome and 32 patients who had a Severe outcome. As shown in Table 4A, only advanced age (OR: 8.0 [95% CI: 3.2, 20.7], p < 0.001), male gender (OR: 3.1 [95% CI: 1.2, 8.7], p = 0.02), and having been diagnosed with obesity (OR: 3.7 [95% CI: 1.5, 9.5], p = 0.004) were significant predictors of a more severe SARS-CoV-2 illness. Given the significant association identified between having obesity and a more severe outcome, an additional penalized multivariable logistic regression model was fit (Table 4B), where more distinct categories were used for diabetes, hypertension (HTN), and obesity. Similarly, in this model, advanced age (OR: 9.4 [95% CI: 3.4, 27.4], p < 0.001) and male gender (OR: 3.2 [95% CI: 1.2, 9.8], p = 0.02) were still significant predictors of a more severe outcome. In addition, compared to not being diagnosed with obesity, all 3 levels of obesity (1: BMI = 30.0 – 34.9; 2: BMI = 35 – 39.9; 3: BMI = 40.0+) were significantly associated or just over the threshold for significance (OR: 3.5, p = 0.05; OR: 5.2, p = 0.03; OR: 13.9, p = 0.01) for increasing the odds of a more severe outcome. While not statistically significant at the 0.05 level, compared to having an HbA1C level < 6, having an HbA1C level of 7.1 – 8.0 (OR: 15.5 [95% CI: 0.8, 244.8], p = 0.06) showed a clinically significant association.

**Table 4A:** Penalized Multivariable Binary Regression Model Results with binary comorbidity categories.

| | $\beta$ coef. | SE | P Value | | AOR | 95% CI LB | 95% CI UB |
| --- | --- | --- | --- | --- | --- | --- | --- |
| INTERCEPT | -2.726 | 1.961 | 0.047 |  | - | - | - |
| AGE GROUP = 65+ | 2.080 | 0.471 | < 0.001 |  | 8.007 | 3.249 | 20.722 |
| GENDER = MALE | 1.117 | 0.497 | 0.016 |  | 3.055 | 1.217 | 8.672 |
| RACE/ETHNICITY = HISPANIC | -0.878 | 2.031 | 0.631 |  | 0.415 | 0.023 | 64.881 |
| RACE/ETHNICITY = MIXED RACE | -1.320 | 2.617 | 0.559 |  | 0.267 | 0.001 | 61.757 |
| RACE/ETHNICITY = NON-HISPANIC BLACK | 0.747 | 2.298 | 0.699 |  | 2.111 | 0.052 | 414.054 |
| RACE/ETHNICITY = NON-HISPANIC WHITE | -1.769 | 1.948 | 0.358 |  | 0.170 | 0.012 | 24.652 |
| DIABETES = YES | 0.380 | 0.525 | 0.469 |  | 1.462 | 0.508 | 3.961 |
| OBESITY = YES | 1.296 | 0.464 | 0.004 |  | 3.655 | 1.500 | 9.517 |
| HISTORY OF HYPERTENSION = YES | 0.553 | 0.443 | 0.212 |  | 1.738 | 0.726 | 4.121 |
| CURRENT SMOKER = YES | -0.280 | 1.042 | 0.780 |  | 0.756 | 0.061 | 4.357 |
| ASTHMA = YES | 1.182 | 0.784 | 0.135 |  | 3.262 | 0.667 | 13.479 |
| COPD = YES | 3.397 | 2.393 | 0.138 |  | 29.878 | 0.186 | 725.447 |

**Table 4B:** Penalized Multivariable Binary Regression Model Results with detailed comorbidity categories.

| | $\beta$ coef. | SE | P Value | | AOR | 95% CI LB | 95% CI UB |
| --- | --- | --- | --- | --- | --- | --- | --- |
| INTERCEPT | -2.772 | 1.967 | 0.045 |  | - | - | - |
| AGE GROUP = 65+ | 2.236 | 0.516 | 0.000 |  | 9.351 | 3.438 | 27.370 |
| GENDER = MALE | 1.162 | 0.518 | 0.018 |  | 3.197 | 1.209 | 9.780 |
| RACE/ETHNICITY = HISPANIC | 0.060 | 2.056 | 0.972 |  | 1.062 | 0.053 | 170.106 |
| RACE/ETHNICITY = MIXED RACE | -2.659 | 2.790 | 0.281 |  | 0.070 | 0.000 | 18.420 |
| RACE/ETHNICITY = NON-HISPANIC BLACK | 0.853 | 2.308 | 0.660 |  | 2.347 | 0.058 | 468.343 |
| RACE/ETHNICITY = NON-HISPANIC WHITE | -1.687 | 1.951 | 0.377 |  | 0.185 | 0.013 | 26.871 |
| DIABETIC.CATEGORY = HBA1C < 7.0 | 1.389 | 0.903 | 0.124 |  | 4.009 | 0.656 | 20.517 |
| DIABETIC.CATEGORY = HBA1C: 7.1 - 8.0 | 2.738 | 1.462 | 0.062 |  | 15.459 | 0.845 | 244.781 |
| DIABETIC.CATEGORY = HBA1C: 8.1 - 9.0 | 0.201 | 1.242 | 0.868 |  | 1.223 | 0.100 | 11.625 |
| DIABETIC.CATEGORY = HBA1C > 9.0 | -0.754 | 1.803 | 0.639 |  | 0.470 | 0.003 | 7.939 |
| HYPERTENSION.CATEGORY = ELEVATED | 0.513 | 0.937 | 0.587 |  | 1.670 | 0.238 | 9.590 |
| HYPERTENSION.CATEGORY = HTN STAGE 1 | 0.437 | 0.651 | 0.504 |  | 1.548 | 0.426 | 5.631 |
| HYPERTENSION.CATEGORY = HTN STAGE 2 | -1.011 | 0.648 | 0.118 |  | 0.364 | 0.094 | 1.285 |
| OBESITY.CATEGORY = BMI: 30.0 – 34.9 | 1.262 | 0.633 | 0.050 |  | 3.533 | 0.998 | 12.641 |
| OBESITY.CATEGORY = BMI: 35.0 – 39.9 | 1.647 | 0.749 | 0.033 |  | 5.189 | 1.147 | 23.908 |
| OBESITY.CATEGORY = BMI: 40.0+ | 2.631 | 0.922 | 0.007 |  | 13.889 | 2.113 | 87.043 |

Coef. = coefficient estimate; SE = Standard Error;
AOR = Adjusted Odds Ratio; CI = Confidence Interval; LB = Lower Bound; UB = Upper Bound

As shown in Supplementary Table 1, only 5.2% patients with no underlying comorbid conditions were in the severe category as compared to patients with one comorbidity; diabetes (16.7%), hypertension (11.8%), or obesity (8.9%). Additionally, when evaluating combinations of the comorbid conditions, 28.1% of patients with hypertension and obesity, followed by 14.3% of patients with diabetes and obesity and 12.5% of patients with diabetes and hypertension, had a more severe outcome. Furthermore, among patients who had all three underlying comorbid conditions (n = 13), 30.8% had more severe illness.

## Discussion

In patients presenting with SARS-CoV-2 infection, there is general consensus that non-modifiable risk factors, older age and male gender, are predictors of severe disease among hospitalized patients^12^. Our findings in the primary care population are consistent; older age and male gender predicted a significantly higher likelihood of severe illness (OR:9 and OR: 3 respectively, p<0.05). Regarding ethnicity, while underrepresented minorities have been identified with more severe outcomes,^13^ our study did not find a statistical significance.

However, among modifiable risk factors, there is greater evidence on the inpatient population, as compared to the ambulatory primary care population, that utilizes comorbidities as predictors of severity of illness. To our knowledge, this is the first study which predicts the progression of illness in the ambulatory population based solely on underlying high risk comorbid conditions, such as those of the components of metabolic syndrome.

We identified a significant association between the number of comorbid conditions and patients with mild (asymptomatic) vs severe (requiring home oxygen or hospitalization) outcomes (Mean (SD): 0.7 (0.9) vs 1.5 (1.1), p< 0.01). For each additional comorbidity, the odds of having a more severe outcome were 2.1 [95% CI: 1.40, 3.10] times higher. This is consistent with inpatient literature, identifying the risk of death in patients with 2-5 underlying comorbid conditions was 2.55 times that of those without^14^. We further evaluated the components of metabolic syndrome, specifically obesity, hypertension, and diabetes, since they have been identified as predictors of more severe illness, due to an underlying proinflammatory state, and increased expression of ACE2 receptors in lung and adipose tissue^15,16,17^. Our study showed that 30.8% of patients with obesity, hypertension, and diabetes together had a severe outcome as compared to 22.2% with two of the three comorbid conditions, and less than 17% with diabetes, hypertension, or obesity alone.

Notably, our findings (Table 4A) show that obesity alone had a fourfold increase on progression to severe illness, especially those with a BMI of 40+ showing a nearly 14 times higher likelihood of severe SARS-CoV-2 infection. This is consistent with Yang et al, where a higher BMI was associated with increased incidence of severe SARS-CoV-2 infection, especially as obesity is related to increased expression of ACE2 and subsequent end organ damage, and respiratory compromise leading to acute respiratory distress syndrome (ARDS) due to increased abdominal pressure limiting chest expansion^18,19,20,21^.

Hypertension and diabetes are among the most common underlying comorbid conditions in patients infected with SARS-CoV-2^22^, with a relative risk of 2.84 and 2.61 respectively^23^, however, to date there is no evidence of whether these are independent risk factors or additive. While our study showed that the presence of hypertension was predictive of more severe illness, it was not a statistically significant independent risk factor (p = 0.21). (Table 4A). Additional analysis evaluating the stages of hypertension (Table 4B) showed that patients who presented in Stage 3 exhibited a lower likelihood of having severe illness (OR [95% CI]: 0.36 [0.09, 1.29]) compared to normotensive patients. Additionally, there was no significant association with disease severity (p = 0.50) in patient prescribed an ACEI or ARB. Our results suggest that while hypertension may play a role in cardiovascular illness, it is not an independent risk factor for acute SARS-CoV-2 related illness and requires further evaluation.

Diabetes, especially with complication, has also been implicated in severity of SARS-CoV2 illness^14^. In our study, hypertension and/or diabetes accounted for 73.6% of the patients, and the prevalence of diabetes alone was 16.7% in the severe category which was higher than hypertension or obesity alone (Supplemental Table 1). While diabetes in general has been well established as a higher risk factor there has been limited evaluation of the stages of diabetes control and its relationship with severe illness. Our analysis shows that patients with a baseline A1C of greater than 6.5 mg/DL (category 2-4) had an increasing likelihood of progression to severe illness. (Table 4B). The increase was only clinically significant for diabetic patients with an A1C between 7.1 - 8.0 mg/dL (category 3), who exhibited a 15.5 times higher likelihood of progression to severe disease (p=0.06). Patients with an A1C > 9.0 had the lowest odds of progression to severe illness (OR [95%CI]: 0.47 [0.00, 7.94], p = 0.64). One possible explanation is that diabetes requires a balance of control^24^ which requires further evaluation as the use of insulin and oral hypoglycemics was not available in our study. Since SARS-CoV-2 has the ability to destroy islets via the angiotensin-converting enzyme 2 (ACE2) receptor, and promote hyperglycemia during acute illness^25^, further analysis evaluating fasting blood sugar will delineate the risk of disease progression among SARS-CoV-2 patients with underlying diabetes.

Numerous studies have identified an association between acute SARS-CoV-2 infection, disease progression and presence of underlying comorbid conditions. As such, there is an increased need to closely monitor patients in the ambulatory setting, both as preventative, prior to infection, and during acute infection with SARS-CoV2. To augment preventative measures, there is a role for primary care physicians in enrolling patients in nutrition and weight loss programs to encourage control of diabetes. Once infected there is a role for closer monitoring of the SARS-CoV-2 infected patients who have additional underlying comorbidities to prevent severe decompensation. This may include augmented monitoring of hypoxia with ambulatory pulse oximetry, improving pulmonary rehab with the use of incentive spirometers, augmented access to point of care glucose monitoring in the ambulatory setting, and increased frequency of tele visits for earlier identification of severe disease progression.

## Limitations

There are several limitations due to the observational and retrospective chart review study design. First, this is a single FQHC in Lakewood, New Jersey with the majority patients identified as Caucasian. Second, there is a small sample size of 150 patients in each arm; however, they were matched by age category and gender in an attempt to help counteract these potential confounders. Third, all the data collected about underlying comorbid conditions was identified based on chart review at the time of presentation for severe illness, and additional follow up questions from the survey may have potentially been affected by patient recall bias. Due to the high volume and large scale of the pandemic, labs were not routinely checked and medication adherence for comorbid conditions was not routinely monitored.

## Conclusion

The progression of SARS-CoV-2 infection can be prognosticated in the ambulatory population by the presence and severity of pre-existing comorbidities. Early identification and risk stratification of these comorbidities will allow clinicians to develop plans for closer monitoring to prevent severe illness.

## Data Availability

All data produced in the present work are contained in the manuscript

## Abbreviations

ARDS: Acute Respiratory Distress Syndrome
COPD: Chronic Obstructive Pulmonary Disease
EMR: Electronic Medical Record
HbA1C: glycosylated hemoglobin
BMI: Body Mass Index

## Acknowledgments

The authors thank Michael Steinberg, MD for contributing to the review of the manuscript from an ambulatory physician perspective and for Yaakov Schwartz, BA for facilitating access to these data. The findings and conclusions in this report are those of the authors and the authors have no funding to disclose. No copyrighted materials were used in this article.

## Author Information

Corresponding Author: Payal Parikh, MD, FACP Rutgers, Robert Wood Johnson Medical School. Clinical Academic Building, 125 Paterson Street, CAB 2320, New Brunswick, NJ 08901. Telephone: 732-325-7254. Twitter: @Payalia320

## Tables

**Supplemental Table 1:** Percentage of Patients with COVID-19 Severity Outcome as a Function of Single and Combination of Underlying Comorbid Conditions.

| Comorbidity Combination, n (Row %) | COVID-19 Severity |  | Total |
| --- | --- | --- | --- |
|  | Mild (1) | Severe (2) |  |
| 0 - NONE | 147 (94.8) | 8 (5.2) | 155 |
| 1 - DIABETES | 5 (83.3) | 1 (16.7) | 6 |
| 1 – HISTORY OF HYPERTENSION | 15 (88.2) | 2 (11.8) | 17 |
| 1 - OBESITY | 51 (91.1) | 5 (8.9) | 56 |
| 2 - DIABETES + HISTORY OF HYPERTENSION | 7 (87.5) | 1 (12.5) | 8 |
| 2 - DIABETES + OBESITY | 12 (85.7) | 2 (14.3) | 14 |
| 2 – HISTORY OF HYPERTENSION + OBESITY | 23 (71.9) | 9 (28.1) | 32 |
| 3 – DIABETES + HISTORY OF HYPERTENSION + OBESITY | 9 (69.2) | 4 (30.8) | 13 |
| Total | 269 | 32 | 301 |

**Supplemental Figure 1:**
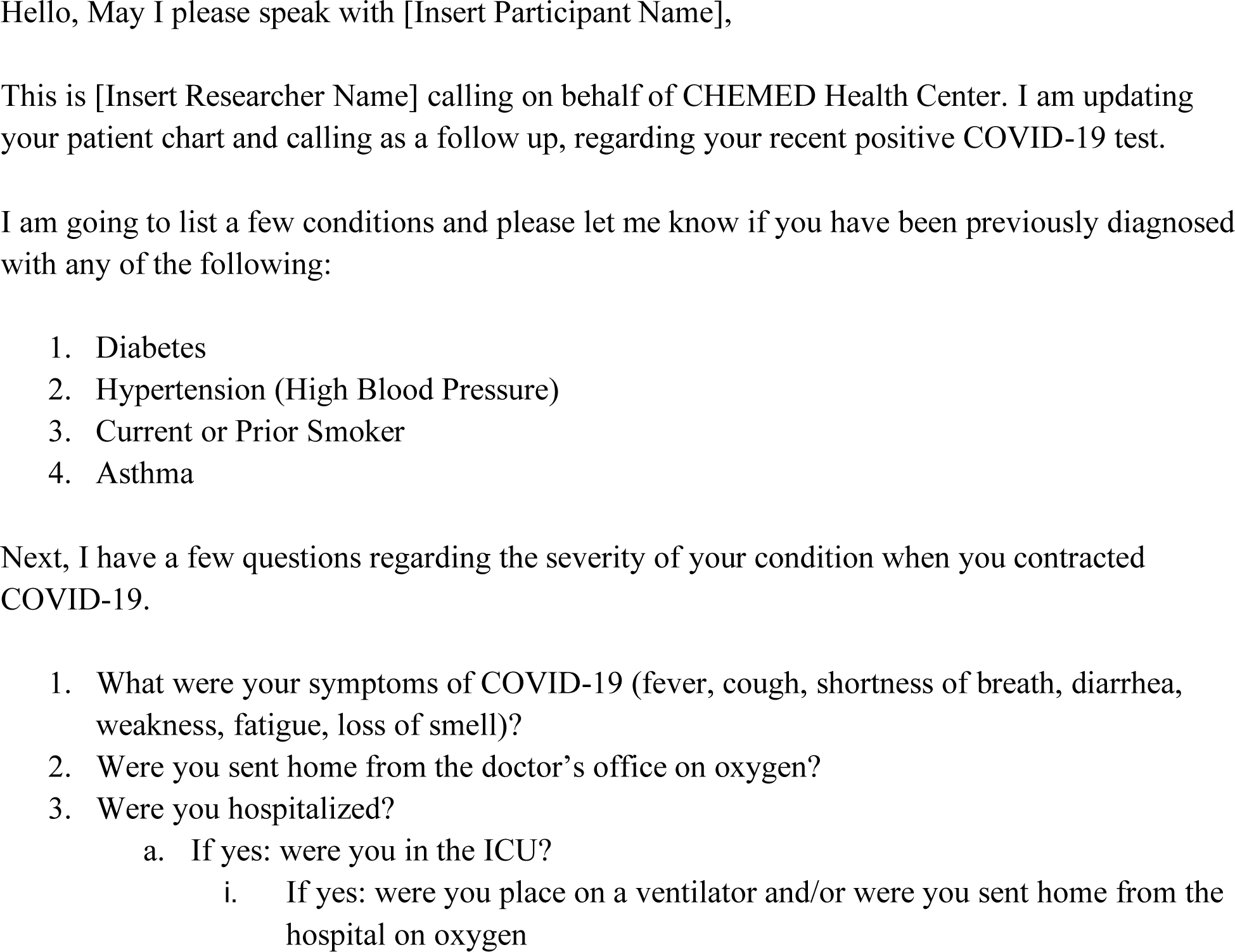
Script for Patient Follow-Up.

